# Building Critical Care Capacity in a Low-Resource Setting: Lessons Learned from Establishing an ICU in Uganda

**DOI:** 10.64898/2026.09.08.26362483

**Authors:** Niels J. Jansen, John Manzi, Sil van den Bersselaar, Lars I. Veldhuis

## Abstract

**Background:** Access to intensive care unit (ICU) services remains severely limited in many low- and middle-income countries (LMICs). Although conceptual frameworks and policy-level recommendations for critical care development exist, real-world experiences describing the practical challenges of starting an ICU in a low-resource setting are scarce.

**Methods:** This mixed-methods descriptive study was conducted at a rural hospital in Uganda. A six-bed ICU was established with capabilities for mechanical ventilation, inotropic support, continuous monitoring, point-of-care blood gas analysis, and piped oxygen. Quantitative data including demographics, diagnoses, and interventions were collected. In addition, qualitative data were collected through structured group discussions to identify key challenges and lessons learned.

**Results:** A total of 45 patients were admitted during the first three months. Key challenges identified included an unstable power supply, procurement delays for essential medications, limited baseline critical care knowledge, cultural barriers to assertive communication, and financial constraints. Continuous training, standardized protocols, and continuous patient monitoring were critical components of successful ICU operation.

**Conclusions:** Establishing a functional ICU in a low-resource setting is possible, but requires extensive planning, continuous training, and robust infrastructure. Simple and affordable interventions, such as continuous monitoring, improved nurse-to-patient ratios, and reliable oxygen supply, had a substantial impact on patient safety. This study provides practical insights that complement existing policy frameworks and inform similar initiatives that strengthen critical care in LMICs.

**Key Points:** 

**Question:** What practical challenges and lessons are encountered when establishing and initiating a six-bed intensive care unit in a rural, low-resource setting?

**Findings:** In this mixed-methods descriptive study of the first 3 months of a newly established ICU in rural Uganda, 45 patients were admitted, while stakeholder discussions identified unstable power supply, procurement delays, limited critical care knowledge, communication barriers, and financial constraints as key implementation challenges. Continuous training, standardized protocols, and continuous patient monitoring were identified as important components of successful ICU operation.

**Meaning:** Establishing critical care capacity in a rural low-resource setting is feasible but requires structured planning, reliable infrastructure, continuous workforce training, and adaptation of ICU systems to local circumstances.

## Introduction

Intensive care units (ICUs) are the cornerstone of modern healthcare systems that provide specialized care for patients with life-threatening conditions and organ failure through continuous monitoring and advanced life-support therapies. ICUs play a vital role in the management of critical illnesses, including sepsis, acute respiratory distress syndrome (ARDS), severe trauma, and complications arising from surgical, medical, or obstetric emergencies. In high-income countries, ICU care is an integral component of standard hospital services supported by the widespread availability of ICU beds, standardized clinical protocols, and well-trained multidisciplinary teams [1,2]. This level of critical care is not universally available. In many low- and middle-income countries (LMICs), access to ICU services is limited or entirely absent due to systemic constraints. In sub-Saharan Africa (SSA), ICU bed availability is often less than 0.5 beds per 100.000 population — with beds mostly concentrated in urban tertiary hospitals, while the region bears a high burden of critical illness due to endemic infectious diseases (e.g., malaria, HIV/AIDS, sepsis), road traffic injuries, obstetric complications, and rising non-communicable diseases [3,4]. This is in high contrast to Europe, where there is an average of 11.5 ICU beds per 100,000 head of the population [1].

Uganda provides a compelling example of these challenges and the ongoing efforts to strengthen ICU services within a constrained health system. As of 2020, Uganda has twelve functioning ICUs of which ten are located in the capital Kampala. Together, they have 55 functional ICU beds for a population of almost 50 million, with most beds located in private or referral hospitals in urban areas. Consequently, access to critical care is highly inequitable, with many district hospitals lacking ICU services [5,6].

Health systems in these settings often lack infrastructure, workforce, technical expertise, and financial resources required to establish and sustain ICU care. Malelelo-Ndou et al. stated that the government is responsible for improving the infrastructure to ensure equitable access to critical care [7]. Common barriers described in multiple studies include insufficient number of ICU beds, shortages of trained physicians and nurses, limited availability of essential medications and monitoring equipment, and unreliable access to electricity and medical oxygen [8–10].

A recent narrative review described the potential difficulties in starting an ICU [11]. However, most of the findings were quite generic. The recommendations made by previous studies are most useful for policymakers at macro level [7–10]. However, real-life practical difficulties in starting and running an ICU have not been well reported. Therefore, this article goes beyond ideas and concepts and describes the process of establishing a six-bed ICU in a rural Ugandan hospital and report the practical challenges, adaptations, and early operational experiences encountered during implementation.

## Methods

### Study design and setting

This mixed-methods descriptive study was performed at St. Francis Hospital Mutolere, Uganda, from October 2024 to June 2025. October 2024 to April 2025 comprised the design and implementation phases, while April to June 2025 represented the early operational phase. St. Francis Hospital Mutolere is a private non-profit church-based hospital with a total of 220 beds. The hospital serves patients from within the Kisoro District and from the neighbouring districts of Rukungiri, Kanungu, Rubanda, and Kabale and approximately 5-10% of the patients come from across the border (Democratic Republic of the Congo and Rwanda). The estimated catchment area is just over 1.000.000 people, and is shared with multiple smaller public health centres and a public district hospital. Historically, Mutolere Hospital has functioned as an unofficial referral centre in the region because of its strong reputation for clinical care. The rural nature of Kisoro District contributes to poor accessibility (physical, political, and financial) and makes it relatively isolated from the rest of the country.

Following careful planning, a well-equipped ICU was established at the Mutolere Hospital. The ICU was designed to provide specialized care for critically ill patients with six beds, including the ability to administer piped oxygen, provide mechanical ventilation, administer inotropic drugs, perform point-of-care blood gas analysis, and ensure continuous patient monitoring by adequately trained personnel.

This study adhered to the STROBE (Strengthening the Reporting of Observational Studies in Epidemiology) informed reporting of the quantitative component.

### Participants and data collection

Data were collected from all patients admitted within the first three months after the opening of the ICU from April 4^th^ to June 4^th^, 2025. Data were extracted from the patients’ charts, vital sign observation records, and medical files after obtaining informed consent from the patient or their next of kin. Collected variables included demographic characteristics, length of ICU stay, discharge diagnosis, Early Warning Score (EWS) at ICU admission, known comorbidities, and the primary reason for admission to the ICU (e.g., mechanical ventilation, circulatory support, or intensive monitoring).

#### Qualitative data

To identify challenges and lessons learned during ICU implementation, structured stakeholder discussions were conducted throughout the planning, implementation, and early operational phases of the project. Participants included hospital management, physicians, nurses, biomedical technicians, and project coordinators involved in ICU development. Detailed notes were recorded during meetings and reviewed by the study team. Recurring challenges, solutions, and implementation experiences were identified through repetitive review of the discussion notes. Findings were grouped into common themes by consensus among the investigators and subsequently organized according to the different phases of ICU implementation.

### Ethical approval

Approval for the study was obtained from the Saint Francis Hospital Mutolere internal ethics review committee, no approval number was provided. The study was carried out in accordance with the principles of the Declaration of Helsinki. Written informed consent was obtained from the patient or their next of kin when the patient was a minor or unconscious. Patients or the public were not engaged in the research planning, conduct, or design.

## Results

During the stakeholder discussions, the experiences and challenges encountered were discussed and organized into three groups: the design and planning phase, the implementation and construction phase, and the early operational phase. These lessons learned are summarized in Table 1, and the implementation timeline is presented in Figure 1.

**Figure 1.**
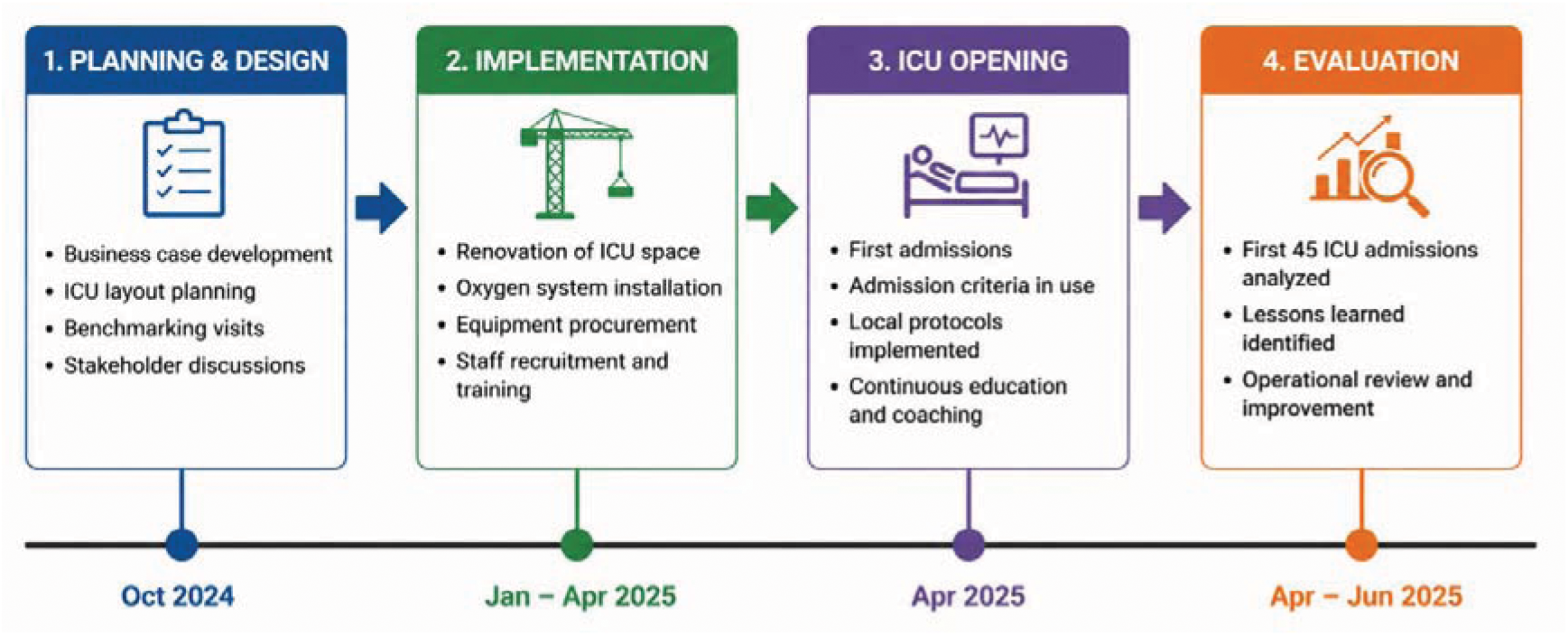
Timeline of the planning, implementation, ICU opening, and evaluation phases of establishing a six-bed ICU in Uganda.

**Table 1.**
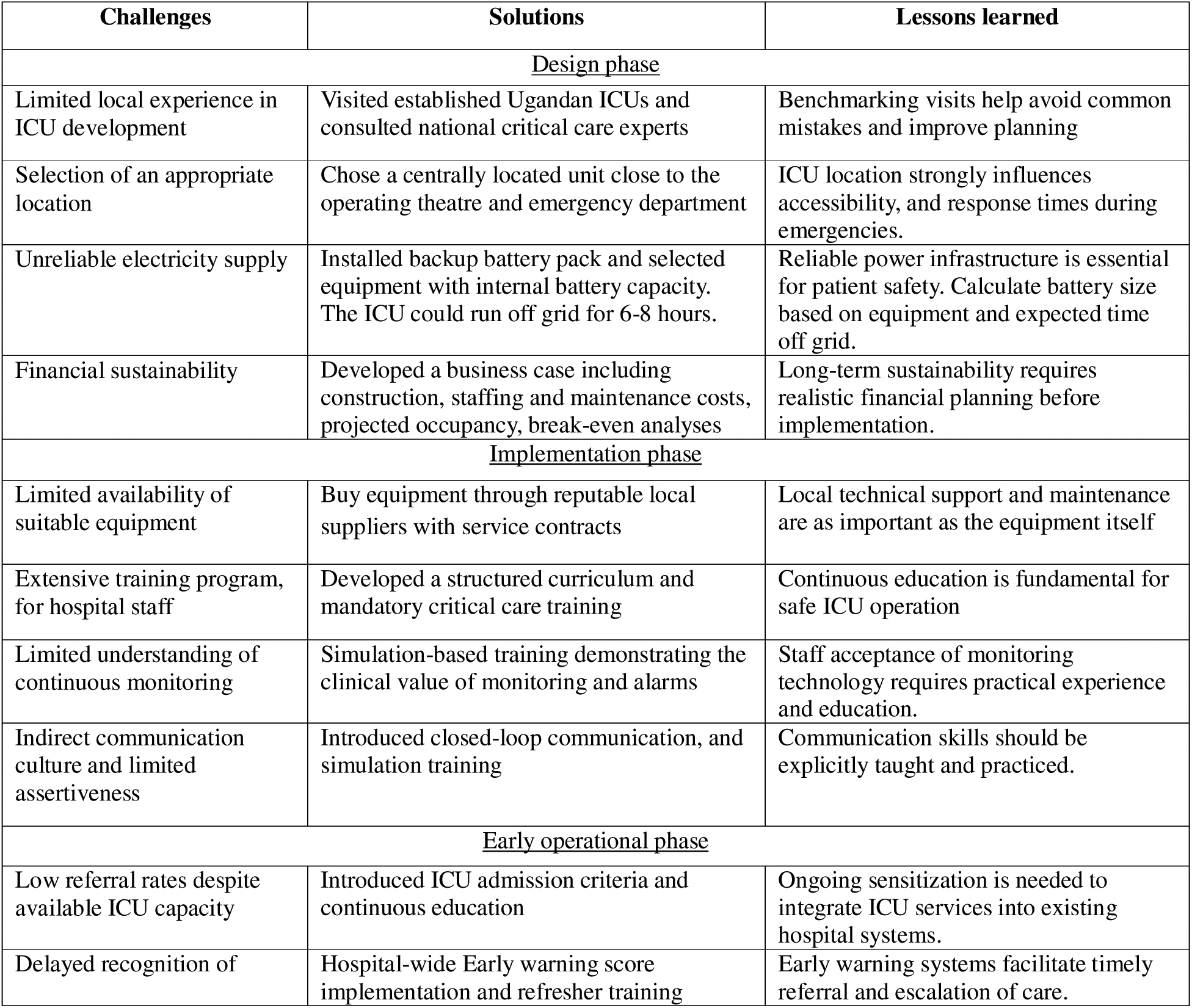
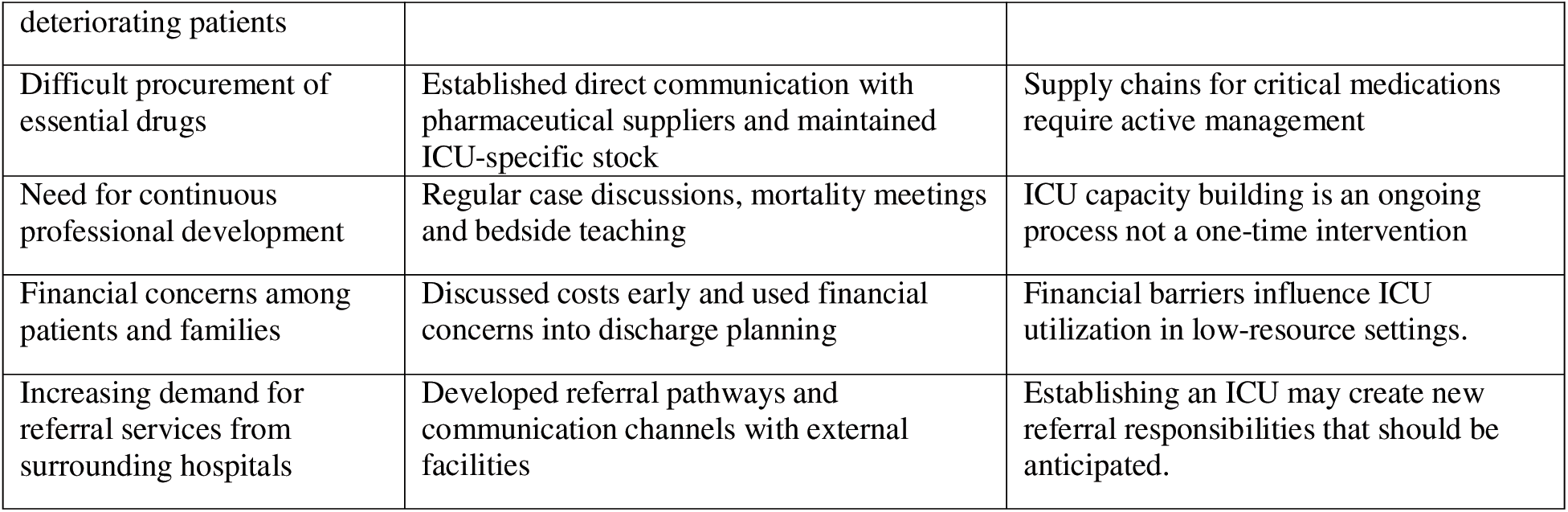
Most valuable lessons learned.

### Design and planning phase

To improve our knowledge, we visited several other hospitals in Uganda that already had an ICU. Three key lessons were identified during this phase.

First, the infrastructure and layout of the ICU should be optimized. This meant that the used space was large enough and allowed safe and easy movement of staff and equipment, especially during emergencies. Monitors showing vital signs should be easily visible to nurses during both day and night. In addition, the ICU should be close to the operating theatre and emergency department, as most of the patients requiring ICU admission are referred from these wards.

Second, the power supply is unstable in many LMICs. Interruptions of the power supply can be life-threatening, especially for patients needing mechanical ventilation or inotropic drugs administered by power supply dependent syringe pumps. Therefore, a reliable backup power system is essential, as is equipment capable of operating for a limited time without an external power source. This may be achieved through internal batteries; however, it is crucial that the equipment continuous to function without rebooting, as a ventilator that shuts down may result in patient harm.

Finally, a robust business case was crucial to ensure that the costs of construction, operation expenses, and the cost per bed per night were all within possible ranges to prevent financial difficulties in the longer run. An overview of the initial investment costs is presented in table 2.

**Table 2.** Investment costs of establishing a six-bed intensive care unit in Uganda.

| Category | Examples | Cost (USD) |
| --- | --- | --- |
| **Infrastructure and renovation** | Building modifications, oxygen piping, security camera's | \$12.975 |
| **Medical equipment** | Monitors, ventilators, infusion pumps, suction devices, arterial blood gas analyser, suction devices, etc. | \$22.397 |
| **Furniture and fixtures** | ICU beds, trolleys, electrical works | \$8.261 |
| **Training and education** | Critical care courses, simulation training, educational materials | \$1.568 |
| **Total implementation cost** | | **\$45.201** |

### Implementation and construction phase

During this phase, the focus was on three main pillars: construction, equipment, and adequate training.

To reduce costs, we aimed to purchase equipment and materials locally and secure maintenance and repair services through local contracts. To prevent equipment failure and ensure reliability, we bought products only from reputable suppliers.

As the management of critically ill patients was new to the hospital, a comprehensive training curriculum was developed for both doctors and nurses. A standardized operating procedures (SOP) manual was created for the frequent reasons for ICU admission and diseases, and was made accessible to all healthcare providers.

To improve awareness and understand the potential of ICU care, we ensured that healthcare providers understood the role of the ICU compared to non-ICU care. The following procedures were implemented; training on the Early Warning Score (EWS) system was provided to all hospital wards, a list of ICU admission criteria was developed and made accessible for all healthcare providers and a five day “Critical Care in Low-Resource setting” course was organized in collaboration with Kabale University hospital. The completion of this course was mandatory for all nurses prior to working in the ICU, ensuring a minimum standard of competency in critical care delivery.

In addition, all newly appointed ICU nurses and doctors participated in two weeks of daily training sessions. These sessions included practical instruction in cardiopulmonary resuscitation (CPR), Crew Resource Management (CRM), and simulation-based scenarios addressing respiratory and hemodynamic instability.

An important observation during this period was communication; culturally, this was more indirect. Direct task allocation and assertive communication were initially challenging. For example, asking for assistance from a colleague was new and difficult to implement. Therefore, these training sessions focused on (closed loop)communication and teamwork principles. Participants reported that this approach was highly beneficial, noting improvements in workflow, teamwork, and patient outcomes.

In addition, nurses initially perceived continuous monitoring and its alarms as disruptive, leading them to silence or turn off the monitors. After personally experiencing how the monitoring and alarm system could be used as a tool to improve patient outcomes during the training sessions, the nurses understood its benefits and incorporated them into daily practice.

### Early operation phase

Despite an extended planning and implementation phase, several lessons emerged during the first three months after opening the ICU.

Many wards have traditionally managed conditions such as acute renal failure, hypertensive emergencies, or diabetic ketoacidosis without ICU involvement. After the establishment of the ICU, the number of ICU admissions initially remained relatively low. Simultaneously, patients were identified in wards with elevated EWS, warranting ICU consultation, or with diagnoses that would have been more appropriately managed in the ICU setting. However, in many cases, the ICU was not consulted, nor were the patients referred to the ICU.

Therefore, additional training sessions were conducted for ward nurses and physicians, focusing on appropriate ICU referral criteria and how to use the EWS. Furthermore, posters outline EWS thresholds, and ICU consultation criteria are displayed in each ward to standardize referral practices. Another important observation is the influence of the patient’s financial situation. After initial stabilization in the ICU, several patients or their families requested early transfer to the general ward because of concerns about hospital costs. In addition, (re)-admission to the ICU in case of deterioration in the ward was a frequent reason for discussion due to financial concerns.

Despite proper planning and good contact with pharmaceutical companies, the challenge was to get the right medication in time. Especially antibiotics and vasopressors such as noradrenaline were difficult to stock. To improve the availability of essential drugs, ICU staff were responsible for their own orders of essential drugs from suppliers in the capital city. By bypassing the hospital procurement system, much bureaucracy was cut out.

Medical personnel highlighted the open-plan layout, lower nurse-to-patient ratio, and availability of continuous patient monitoring as major improvements. This new environment greatly increased healthcare workers’ knowledge and motivation.

### Patient characteristics of the ICU admissions

In the first three months after the opening of the ICU, 45 patients were admitted. Of these, 82% (n=37) were male and 18% were female (n=8). Most patients were adults (91%, n=41), and only 4 were aged under 16 years (9%). All patients were Ugandan nationals, with a mean age of 46.4 years (SD=21.2 years). The majority (40%) were admitted directly from the operating theatre (OR) and 31% were admitted from the surgical ward. In total, six patients were transferred from another hospital to the ICU for its expertise and capabilities. Most patients had problems related to surgery (62%), followed by medical, obstetrics and gynaecology, and paediatrics (29%, 7%, and 2%, respectively). The most common indication for ICU admission was post-operative care requiring intensive monitoring, post-operative ventilation, or supplemental oxygen therapy (36%), followed by altered mental status (29%), and sepsis/shock (20%). All patients tested negative for HIV and the majority (71%) had no comorbidities. The average length of stay in the ICU was 2.5 days (SD = 1.8 days) and the mean EWS on admission was 8.6 (SD = 3.9), with 58% having an EWS >7 on admission. See table 3.

**Table 3.** Baseline characteristics of study participants.

| Characteristics | All patients (n=45) |
| --- | --- |
| Age in years, mean (SD) | 46.4 ± 21.2 |
| Adult | 41 (91%) |
| Male | 37 (82%) |
| Source of admission |  |
| Ward | 21 (46.7%) |
| Operating theatre | 18 (40%) |
| Other hospital referral | 6 (13%) |
| Admission specialism |  |
| Surgery | 28 (62%) |
| Internal medicine | 13 (29%) |
| Obstetrics and gynaecology | 3 (7%) |
| Paediatrics | 1 (2%) |
| No comorbidities | 32 (71%) |
| HIV negative | 45 (100%) |
| Indication for ICU admission |  |
| Post-operative care | 16 (36%) |
| General surgery | 8 (50%) |
| Neuro surgery | 5 (31%) |
| Others | 3 (19%) |
| Altered mental status | 13 (29%) |
| Sepsis/shock | 9 (20%) |
| Respiratory failure | 4 (9%) |
| Renal failure | 2 (4%) |
| Hypertensive emergency | 1 (2%) |
| EWS on admission | 8.6 ± 3.9 |
| EWS>7 on admission | 26 (58%) |
| Length of stay in ICU (days) | 2.5 ± 1.8 |
| Interventions |  |
| Mechanical ventilation | 4 (9%) |
| Inotropic drugs | 8 (18%) |
| Blood transfusion | 23 (51%) |
| Arterial blood gas analysis | 9 (20%) |

### Interventions and treatments during ICU stay

Of the admitted patients, four (9%) needed mechanical ventilation, with an average of 1.5 days on the ventilator. About one in five patients (18%) required inotropic drugs. More than half (51%) needed at least one blood transfusion, and the transfused patients received 2.4 units per patient on average (Table 3).

## Discussion

Establishing an ICU in a low-resource setting came with a host of challenges —some expected, others only revealed through experience. Financing remains one of the most persistent barriers to critical care development in sub-Saharan Africa. Previous studies have identified limited healthcare expenditure, dependence on donor funding, and competing health priorities as major obstacles to ICU expansion [11]. While the initial investment for our ICU was obtained through crowdfunding, long-term sustainability required development of a detailed business case and realistic cost projections. Our experience suggests that financial planning should begin before construction starts and should include projected occupancy rates, maintenance costs, and staff expenditures. Failure to address sustainability may result in well-equipped units becoming non-functional after initial funding is exhausted. With a structured approach, proper planning, and dedication, it is possible to realize a functional 6 bed ICU in a rural area of a low-income country.

In a narrative review from 2023, recommendations were made based on the WHO framework, which classifies health systems into six core components: (1) service delivery, (2) health workforce, (3) health information systems, (4) access to essential medicines, (5) financing, and (6) leadership/governance [11]. We agree with most of Spencers’ review, although most of their findings are quite generic. For example, insufficient critical care beds, understaffed and undertrained staff, large differences between urban and rural areas, poor access to equipment and drugs, and financial difficulties. The recommendations made by Spencer et al. are most useful for policymakers at macro level. Our study is of additional value because it provides real-life experiences and lessons learned from starting an ICU in a low-resource setting. Therefore, it goes beyond ideas and concepts.

Training emerged as one of the most important determinants of successful ICU implementation. Although infrastructure and equipment are often emphasized as critical care services, our experience suggests that workforce development may be equally important. Similar challenges have been described in other African settings where limited exposure to critical care principles results in delayed recognition of deterioration and reduced quality of care [5, 7]. In our setting, signs of clinical deterioration were sometimes missed or acted on too late. Decisions on whether a patient needed to be reviewed often came down to guesswork or habits, rather than objective assessment. After the introduction of EWS, standardized ICU admission criteria, and a critical care training the decision to admit to the ICU became more standardized and uniform, and the threshold to call for help was thereafter reduced. These interventions required relatively limited financial investment but had a substantial impact on clinical decision-making.

Given that the mean EWS at ICU admission was 8.6, there remains room for improvement in ensuring that deteriorating patients are admitted to the ICU in time. Therefore, additional education is warranted for nurses and doctors at the wards. Conditions such as sepsis, which require fast recognition and coordinated management, were often identified late due to lack of training in recognizing early signs of critical illness. The development of locally adapted protocols proved essential for maintaining consistency of care. Protocols not only standardized clinical practice but also served as educational tools for newly trained staff. Importantly, protocols developed in high-income settings are not always applicable in resource-limited environments because of differences in staffing, diagnostics, medication availability, and infrastructure. Therefore, adaptation to local realities is crucial. Our findings support the concept that protocol development should be viewed as a core component of ICU implementation rather than an optional addition.

Interestingly, the interventions perceived to have the greatest impact on patient safety were not the most technologically advanced. Continuous monitoring, piped oxygen, improved nurse-to-patient ratios, and enhanced patient visibility within an open ward design were considered most influential. These interventions are relatively easy and affordable.

Several limitations should be acknowledged. First, this is a single-center experience from a rural hospital in Uganda. Second, the evaluation period was relatively short and included only the first 45 ICU admissions. Finally, our setting might not always be generalizable to all LMIC’s because of cultural, financial, disease prevalence, or infrastructure differences. Nevertheless, the study provides practical implementation insights that are rarely reported in the literature. And therefore, can help to understand the strengths and pitfalls of our setting and, therefore, still provide valuable lessons for others.

## Conclusion

Overall, the process of setting up the ICU exposed a deep need for structured systems: clear admission criteria, standardized training, reliable supplies, consistent protocols, and better communication across facilities. Despite these challenges, critical care capacity building in LMIC is essential, especially in rural areas. We believe that our experience can help to understand the strengths and pitfalls of our ICU and therefore, provide valuable lessons for others. Furthermore, we are willing to share our financial planning documents, business case, procurement strategies, protocol book, and training materials with institutions interested in developing ICU services in similar settings.

Future research should evaluate the long-term sustainability and impact of newly established ICUs in low-resource settings. Important areas include patient outcomes, cost-effectiveness, staff retention, and the durability of training programs. In addition, studies should assess whether interventions introduced during ICU implementation, such as Early Warning Scores, standardized admission criteria, and protocol-based care, lead to earlier recognition of critical illness and improved patient outcomes. Multi-center studies would help determine the generalizability of these findings to other rural hospitals in sub-Saharan Africa.

## Data Availability

All data produced in the present study are available upon reasonable request to the authors

## Acknowledgments

We would like to thank the staff of St. Francis Hospital Mutolere (Uganda) for helping in every possible way to make this research possible. Special thanks goes to Nsengiyumva Aloiz (nurse in- charge surgical ward and ICU), Kabale University and ICU for their collaboration in training and Arthur Kwizera (Associate Professor of Anaesthesia and Intensive Care, Makerere University) for fruitful discussions during the planning phase.

## Notes

Conflict of interest: No potential conflict of interest or funding was reported by the author(s).

### Competing Interest Statement

The authors have declared no competing interest.

### Author Declarations

Approval for the study was obtained from the Saint Francis Hospital Mutolere internal ethics review committee.

## References

[1] Rhodes A, Ferdinande P, Flaatten H, Guidet B, Metnitz PG, Moreno RP. The variability of critical care bed numbers in Europe. Intensive Care Med 2012;38:1647–53. 10.1007/s00134-012-2627-8.

[2] Halpern NA, Tan KS. United States Resource Availability for COVID-19. Society of Critical Care Medicine 2020.

[3] Murthy S, Leligdowicz A, Adhikari NKJ. Intensive care unit capacity in low-income countries: A systematic review. PLoS One 2015;10. 10.1371/journal.pone.0116949.

[4] Craig J, Kalanxhi E, Hauck S. National estimates of critical care capacity in 54 African countries. MedRxiv 2020. 10.1101/2020.05.13.20100727.

[5] Atumanya P, Sendagire C, Wabule A, Mukisa J, Ssemogerere L, Kwizera A, et al. Assessment of the current capacity of intensive care units in Uganda; A descriptive study. J Crit Care 2020;55:95–9. 10.1016/j.jcrc.2019.10.019.

[6] Atumanya P, Agaba PK, Mukisa J, Nakibuuka J, Kwizera A, Sendagire C. Characteristics and outcomes of patients admitted to intensive care units in Uganda: a descriptive nationwide multicentre prospective study. Sci Rep 2024;14. 10.1038/s41598-024-59031-5.

[7] Malelelo-Ndou H, Ramathuba DU, Netshisaulu KG. Challenges experienced by health care professionals working in resource-poor intensive care settings in the Limpopo province of South Africa. Curationis 2019:1–8. 10.4102/curationis.v42i1.1921.

[8] Fowler RA, Adhikari NKJ, Bhagwanjee S. Clinical review: Critical care in the global context - Disparities in burden of illness, access, and economics. Crit Care 2008;12. 10.1186/cc6984.

[9] Baelani I, Jochberger S, Laimer T, Otieno D, Kabutu J, Wilson I, et al. Availability of critical care resources to treat patients with severe sepsis or septic shock in Africa: A self-reported, continent-wide survey of anaesthesia providers. Crit Care 2011;15. 10.1186/cc9410.

[10] Papali A, Adhikari NKJ, Diaz J V., Dondorp AM, Dünser MW, Jacob ST, et al. Infrastructure and organization of adult intensive care units in resource-limited settings. Sepsis Management in Resource-limited Settings, Springer International Publishing; 2019, p. 31–68. 10.1007/978-3-030-03143-5_3.

[11] Spencer SA, Adipa FE, Baker T, Crawford AM, Dark P, Dula D, et al. A health systems approach to critical care delivery in low-resource settings: a narrative review. Intensive Care Med 2023;49:772–84. 10.1007/s00134-023-07136-2.

